# Radiological Phenotypes of Acute Brain Injury During Extracorporeal Membrane Oxygenation on Ultra-Low-Field Portable MRI

**DOI:** 10.64898/2026.10.21.26366184

**Authors:** Kotaro Noda, Yaman B. Ahmed, Hyun Yi (Jacqualine) Woo, Shivalika Khanduja, Yipeng Wang, Zoe Soulé, Madison Malfitano, Siyu Wang, Bo Soo Kim, Christopher Wilcox, Aaron M Gusdon, Glenn J.R. Whitman, Sung-Min Cho

## Abstract

**Background:** Despite the clinical importance of acute brain injury (ABI) during extracorporeal membrane oxygenation (ECMO), the mechanism of ECMO-associated ABI remains poorly understood, partly because conventional magnetic resonance imaging (MRI) is incompatible with ECMO devices. Ultra-low-field portable MRI (ULF-pMRI) enables high-quality bedside neuroimaging during ECMO. We aimed to characterize imaging phenotypes of ABI and explore mechanisms underlying lesion distribution across ECMO configurations.

**Methods:** In this multicenter retrospective observational study, adults receiving ECMO who underwent bedside ULF-pMRI at Johns Hopkins University and University of Texas–Houston from December 2021 to May 2026 were included. ABI was defined as ischemic stroke (IS), intraparenchymal hemorrhage (IPH), subarachnoid hemorrhage (SAH), or subdural hematoma (SDH). Lesions were characterized according to vascular territory, cortical/subcortical lesion versus deep white matter (WM)/basal ganglia (BG) involvement, laterality, lesion burden, and cerebral small-vessel disease (SVD) using Fazekas scale. Associations between ECMO configuration (central venoarterial [VA], peripheral VA, and venovenous [VV]), echocardiographic findings, and ECMO parameters were examined.

**Results:** Among 70 patients analyzed (55.9 ± 16.2 years; 54.3% male), 29 (41.4%) had ABI. IS was the predominant type of ABI (82.8%), with most strokes affecting the frontal lobe (48.3%). All the central VA-ECMO patients with ABI demonstrated predominantly cortical/subcortical lesions, whereas deep WM/BG involvement was more frequent in peripheral VA-ECMO (53.8%). Peripheral VA-ECMO demonstrated left hemispheric predominance (85% vs. 15% right-sided, *P*=0.002), and deep WM/BG lesions in peripheral were associated with higher ECMO pump flow and lower pulse pressure. Among VV-ECMO patients, bilateral lesions were more common (62.5%), with a high prevalence of deep WM/BG involvement (62.5%) and SVD (75%).

**Conclusions:** ULF-pMRI revealed distinct radiological phenotypes of ABI across ECMO configurations; cortical/subcortical lesions in central VA-ECMO, left-sided predominance in peripheral VA-ECMO and frequent deep WM/BG in VV-ECMO. These findings generate mechanistic hypotheses regarding configuration-specific pathways of ABI and may inform future strategies for risk stratification, prevention and monitoring.

## Introduction

Extracorporeal Membrane Oxygenation (ECMO) is an essential life-support therapy for patients with refractory cardiac and respiratory failure. Approximately 30% of venoarterial (VA)-ECMO patients experience acute brain injury (ABI), including ischemic stroke (IS) and intracranial hemorrhage (ICH) during ECMO support, resulting in a two-to threefold increase in mortality.^1–3^ Despite advances in ECMO technology and patient survival, the underlying mechanisms of ABI remain poorly understood, limiting the development of targeted preventive strategies.

Our previous studies have suggested that higher ECMO pump flow and lower pulse pressure may increase the risk of ABI reflecting endothelial cell injury due to nonpulsatile flow generated by ECMO.^4,5^ In a porcine model, we further demonstrated that rapid reduction of partial pressure of carbon dioxide (PaCO_2_) during ECMO initiation induces cerebral vasoconstriction and impairs cerebral autoregulation.^6^ In addition to these mechanisms related to microvascular dysfunction, hemostatic instability induced by the extracorporeal circuit may contribute to both thrombus formation and bleeding.^7–9^ These observations suggest that multiple pathophysiologic pathways, including embolism, microvascular dysfunction, and hemostatic instability, may contribute to ECMO-associated ABI. However, the relative importance of these mechanisms remains unknown.

Importantly, these mechanisms are unlikely to contribute equally across ECMO configurations. Our previous clinical study and systematic review demonstrated that IS occurs more frequently during VA-ECMO, particularly central VA-ECMO, than during venovenous (VV)-ECMO, while the prevalence of ICH was relatively similar between VA and VV-ECMO.^10,11^ These observations suggest that distinct pathophysiologic pathways may predominate according to ECMO configuration, resulting in different imaging phenotypes of ABI. Whether these configuration-specific mechanisms produce distinct neuroanatomical patterns of brain injury has not been systematically investigated.

The anatomical distribution and lesion pattern of brain injury may provide important insights into the underlying mechanism. For example, cortical and territorial infarcts are typically associated with embolic mechanisms, whereas deep white matter (WM) and basal ganglia lesions may reflect cerebral small-vessel vulnerability, microvascular dysfunction, or hypoperfusion.^12^ Despite its potential mechanistic importance, detailed characterization of ABI topography during ECMO has been limited, since conventional magnetic resonance imaging (MRI), the reference standard for ischemic ABI, is incompatible with extracorporeal life support device due to the safety concerns, such as device migration and malfunction.^13,14^

Recently, we demonstrated the utility of ultra–low-field (ULF; 0.064-T) portable magnetic resonance imaging (pMRI) for the detection of ABI during ECMO.^14^ ULF-pMRI was safely performed at the bedside and detected ABI approximately twice as often as computed tomography (CT). In our multicenter study, we used ULF-pMRI to characterize the imaging phenotypes of ABI during ECMO by evaluating lesion location, vascular territory, laterality, lesion burden, and preexisting cerebral small-vessel disease across central VA-, peripheral VA-, and VV-ECMO. We further examined associations between these imaging phenotypes and ECMO-related physiological and anatomical factors, including pump flow, pulse pressure, and bovine aortic arch anatomy. We hypothesized that distinct ECMO configurations are associated with distinct imaging phenotypes of ABI, reflecting differences in the underlying pathophysiologic mechanisms.

## Methods

### Study design and patient selection

This is a multicenter, retrospective, observational study at Johns Hopkins Hospital and the University of Texas Health Science Cetner at Houston (UTHealth). Adult patients (≥18 years) who received ECMO support and underwent bedside ultra-low-field portable MRI (ULF-pMRI) between December 2021 and May 2026 were included. The detailed study protocol of ULF-pMRI examination was described in our previous work.^14^

Patients were excluded if (1) image quality was inadequate for interpretation because of severe artifacts or poor resolution, (2) ABI occurred before ULF-pMRI, or (3) severe hypoxic-ischemic brain injury (HIBI) precluded detailed assessment of the focal distribution of ECMO-associated ABI. Clinical information and neuroimaging in medical records and ULF-pMRI data stored on secure servers in Hyperfine were investigated. This study was approved by the institutional review boards of Johns Hopkins Medicine (IRB00285716, and IRB00436424) and University of Texas–Houston (HSC-MS-22-0608) following the Declaration of Helsinki. Informed consent was obtained from a legally authorized representative because enrolled patients could not provide consent, and consent forms were stored in the secure file.

### Radiological evaluation of ABI

ABI was defined as IS, intraparenchymal hemorrhage (IPH), subarachnoid hemorrhage (SAH), or subdural hematoma (SDH) based on ULF-pMRI findings. When available, findings from conventional MRI or computed tomography (CT) obtained during the same hospitalization were incorporated to support lesion classification. We assessed the anatomical location of ABI. The lesions were categized according to their anatomical location as involving the frontal, parietal, temporal, and occipital lobes and the insula, hippocampus, cerebellar hemisphere, corona radiata, anterior limb, genu, and posterior limb of the internal capsule, caudate nucleus, putamen, globus pallidus, thalamus, corpus callosum and brainstem. Furthermore, those lesions were grouped into cortical/subcortical lesions and deep WM/basal ganglion (BG) regions based on their anatomical distribution.

In parallel, the vascular territory of ABI was classified as anterior cerebral artery (ACA), middle cerebral artery (MCA), posterior cerebral artery (PCA), vertebral artery (VA), and basilar artery (BA). Deep lesions were defined as involvement of the deep white matter or basal ganglia, including the corona radiata, internal capsule, caudate nucleus, putamen, globus pallidus, thalamus, corpus callosum, and brainstem. Non-parenchymal hemorrhage (SAH and SDH) was assigned to its associated cerebral hemisphere and lobe for regional analyses. Furthermore, the laterality of each ABI lesion was classified as left-side, right-side, or bilateral, without assigning laterality to brainstem and corpus callosum. Lesion burden was not defined as volume but rather quantified as the number of anatomical regions involved.

Preexisting cerebral small-vessel disease (SVD) was assessed using periventricular hyperintensity (PVH), and deep and subcortical white matter hyperintensity (DSWMH) according to the Fazekas scale.^15–17^ SVD was defined as Fazekas grade ≥1 in either PVH or DSWMH.^18^ Cerebral microbleeds and lacunes were not systematically evaluated because susceptibility-weighted imaging was not included in the standard ULF-pMRI protocol. All images were reviewed by a board-certified neurologist. When lesion classification was uncertain, a second board-certified neurologist independently reviewed the images, and the final diagnosis was established by consensus.

### The Assessment of Bovine Arch Anatomy

To evaluate the potential influence of aortic arch anatomy on the distribution and laterality of ABI, available chest CT or CT angiography studies were reviewed to classify aortic arch branching patterns as either standard or bovine arch. A bovine arch was defined as a common origin of the brachiocephalic artery and left common carotid artery, or the left common carotid artery arising directly from the brachiocephalic artery.^19,20^

### Pathophysiological Variables

To investigate potential mechanisms underlying ECMO-associated ABI, we evaluated embolic and hemodynamic factors. Potential embolic sources were identified from transthoracic or transesophageal echocardiography and included the presence of patent foramen ovale (PFO), left ventricle thrombus, and severely impaired ejection fraction (EF) (<30%).^1,21^ The presence of ECMO circuit clot before ULF-pMRI testing was also investigated as a potential embolic source.

Hemodynamic variables include ECMO pump flow rate and pulse pressure (systolic blood pressure minus diastolic blood pressure) after 24 hours, arterial carbon dioxide change (ΔPaCO_2_ = PaCO_2_ at 24 hours after cannulation - PaCO_2_ at cannulation), and partial arterial oxygen (PaO2) level within 6 hours.^1,4,5,22^ Given the reported associations between these physiological perturbations, impaired cerebrovascular autoregulation, and cerebral microvascular injury, we explored their relationships with deep white matter and basal ganglia lesions, which were considered imaging markers of a potential microvascular phenotype.^16^

### Statistical analysis

Continuous variables were summarized as mean ± standard deviation (SD) or median with interquartile range (IQR), as appropriate based on their distribution. Categorical variables were expressed as frequencies and percentages. Baseline characteristics and imaging findings were compared between patients with and without ABI, as well as among ECMO configurations (central VA, peripheral VA, and VV). Differences in categorical variables were assessed using the chi-square test or Fisher’s exact test, as appropriate. Continuous variables were compared using the Student’s t test for normally distributed data or the Mann-Whitney U test for non-normally distributed data. Comparisons among more than two groups were performed using one-way analysis of variance (ANOVA) or the Kruskal-Wallis test, as appropriate.

Lesion characteristics, including vascular territory, anatomical location, laterality, lesion burden, and preexisting small-vessel disease burden, were analyzed descriptively and compared across ECMO configurations. Exploratory analyses were performed to evaluate associations between imaging phenotypes and physiological variables, including ECMO pump flow, pulse pressure, ΔPaCO_2_, echocardiographic findings, ECMO circuit thrombus, and bovine aortic arch anatomy. Given the exploratory nature of this study and the relatively small sample size, no adjustment for multiple comparisons was performed, and all findings should be considered hypothesis-generating. All statistical tests were two-sided, and a P value <0.05 was considered statistically significant. Statistical analyses were performed using R software, version 4.5.1 (R Foundation for Statistical Computing, Vienna, Austria).

## Results

### Patient characteristics

Between December 2021 and May 2026, 75 patients receiving ECMO underwent bedside ULF-pMRI. Two patients with severe diffuse HIBI and brain edema were excluded because focal ECMO-associated ABI could not be reliably assessed. One patient was excluded because of poor image quality and two because of incomplete clinical data. The final cohort included 70 patients (Johns Hopkins Hospital, n=56; UTHealth, n=14) (**Figure S1**).

The mean age was 55.9 ± 16.2 years, and 54.3% were male. Peripheral VA-ECMO was the most common configuration (47.1%), followed by VV-ECMO (34.1%) and central VA-ECMO (18.6%) (Table 1). The mean duration of ECMO support was 15.3 ± 17.3 days (VA; 10.2 ± 8.9 days, VV; 25.1 ± 24.4 days), and ULF-pMRI was performed at a mean of 4.3 ± 5.7 days after cannulation.

**Table 1.** Patient characteristics.

| Variable | All (n=70) | Non-ABI (n=41) | ABI (n=29) | *P* |
| --- | --- | --- | --- | --- |
| Age, mean (SD) | 55.9 (16.2) | 55.6 (17.2) | 56.2 (14.9) | 0.871 |
| Male sex | 38 (54.3%) | 25 (61.0%) | 13 (44.8%) | 0.275 |
| ECMO mode |  |  |  |  |
| Central VA | 13 (18.6%) | 5 (12.2%) | 8 (27.6%) |  |
| Peripheral VA | 33 (47.1%) | 20 (48.8%) | 13 (44.8%) |  |
| VV | 24 (34.1%) | 16 (39.0%) | 8 (27.6%) |  |
| IS history | 7 (10.0%) | 2 (5.0%) | 5 (17.2%) | 0.122 |
| ICH history | 3 (4.3%) | 1 (2.5%) | 2 (6.9%) | 0.568 |
| Hypertension | 37 (53.6%) | 24 (58.5%) | 13 (46.4%) | 0.514 |
| Hyperlipidemia | 28 (40.6%) | 17 (42.5%) | 11 (37.9%) | 0.894 |
| Diabetes | 22 (31.9%) | 14 (35.0%) | 8 (27.6%) | 0.696 |
| CHF | 15 (21.7%) | 9 (22.5%) | 6 (20.7%) | 0.99 |
| CKD | 18 (26.1%) | 14 (35.0%) | 4 (13.8%) | 0.089 |
| Atrial fibrillation | 12 (17.4%) | 9 (22.5%) | 3 (10.3%) | 0.321 |
| Duration of ECMO support, mean (SD) | 15.3 (17.3) | 14.5 (15.4) | 16.5 (19.9) | 0.661 |
| Cannulation to ULF-pMRI testing, days, mean (SD) | 4.3 (5.7) | 3.1 (2.9) | 5.9 (7.9) | 0.078 |

Abbreviations: ABI; Acute brain injury, SD; Standard deviation, ECMO; Extracorporeal membrane oxygenation, VA; Venoarterial, VV; Venovenous; IS; Ischemic stroke, ICH; Intracranial hemorrhage, CHF; Chronic heart failure, CKD; Chronic kidney disease, ULF-pMRI; Ultra-low-field portable magnetic resonance imaging.

ABI was identified in 29 of 70 patients (41.4%), including 21 (45.7%) receiving VA-ECMO and 8 (33.3%) receiving VV-ECMO. Ischemic stroke was the predominant ABI subtype (24/29, 82.8%), whereas SAH, SDH, and IPH occurred less frequently (**Table 2**). Mixed injury patterns were common, including concurrent IS and SAH (n=3) or IS and IPH (n=1). More than half of patients demonstrated multifocal lesions (VA; 11/21, VV; 5/8).

**Table 2.** The subtype of ABI during ECMO.

| ABI subtype | All (n=29) | VA-ECMO<br>(n=21) | VV-ECMO<br>(n=8) |
| --- | --- | --- | --- |
| IS | 24 (82.8%) | 18 (85.7%) | 6 (75%) |
| IPH | 1 (3.4%) | 0 | 1 (12.5%) |
| SDH | 3 (10.3%) | 2 (6.9%) | 1 (12.5%) |
| SAH | 5 (17.2%) | 4 (13.8%) | 1 (12.5%) |
| Combination<br>pattern |  |  |  |
| IS+SAH | 3 (10.3%) | 3 (14.3%) | 0 |
| IS+ICH | 1 (3.4%) | 0 | 1 (12.5%) |

Abbreviations: ABI; Acute brain injury, VA; Venoarterial, VV; Venovenous; ECMO;
Extracorporeal membrane oxygenation, IS; Ischemic stroke, ICH; Intracranial hemorrhage,
SDH; Subdural hematoma, SAH; Subarachnoid hemorrhage.

### Distinct Imaging Phenotypes Across ECMO Configurations

The frontal lobe was the most frequently involved region (48.3%), followed by the parietal, temporal, and occipital lobes, whereas brainstem lesions were uncommon (**Figure 1A**). Consistent with this distribution, the ACA (48.3%) and MCA (44.8%) territories were most commonly involved, while vertebrobasilar infarction was rare (**Figure 1B**).

**Figure 1:**
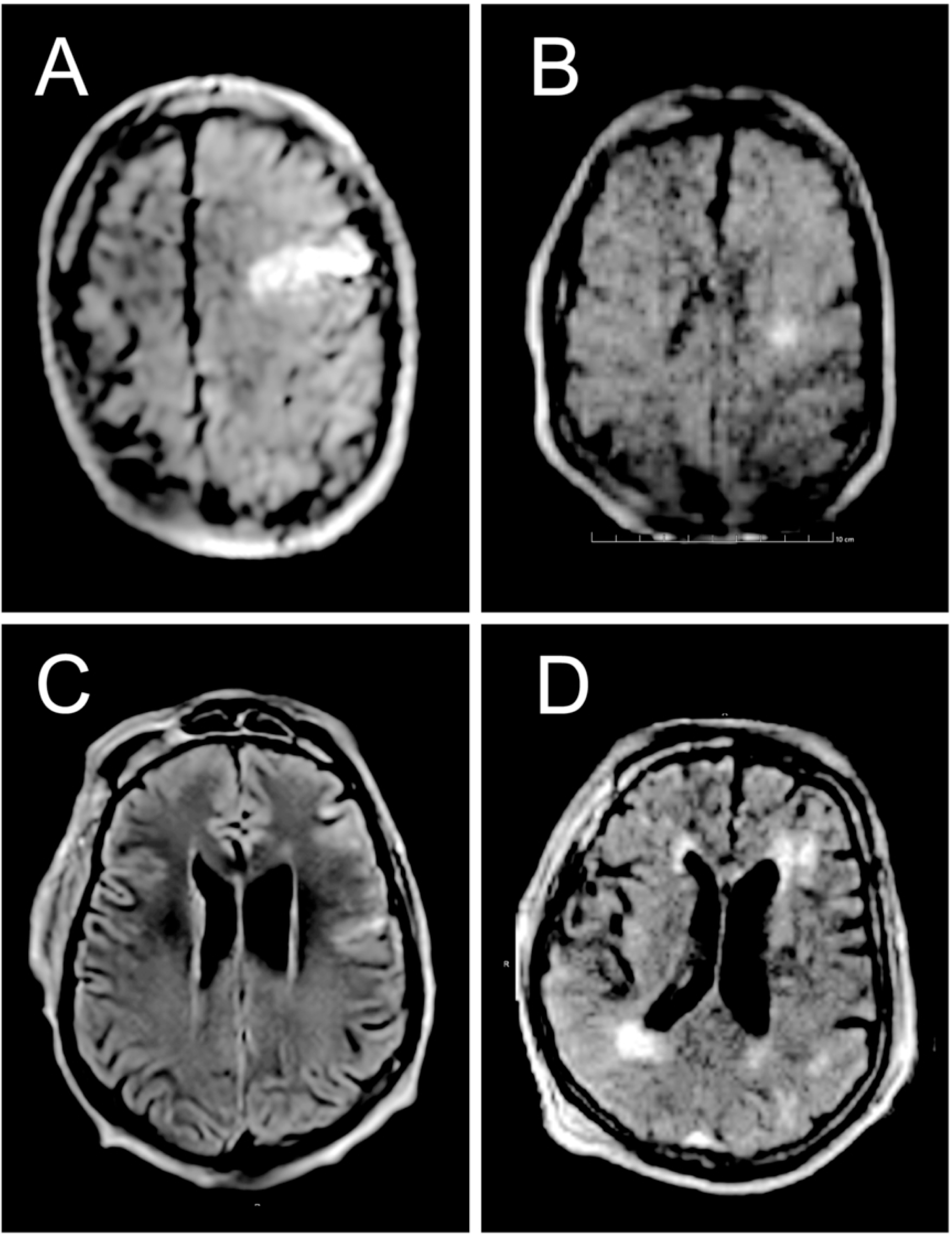
Representative ultra-low-field portable magnetic resonance image findings of acute brain injury. Diffusion weighted imaging (DWI) showed a territorial ischemic lesion on left frontal lobe (A) and perforator infarction involving deep white matter (B). Subarachnoid hemorrhage was found within left frontal lobe sulcus in fluid-attenuated inversion recovery (FLAIR) (C). Grade 2 of periventricular hyperintensity, and grade 2 of deep and subcortical white matter hyperintensity were also observed on FLAIR (D).

**Figure 2:**
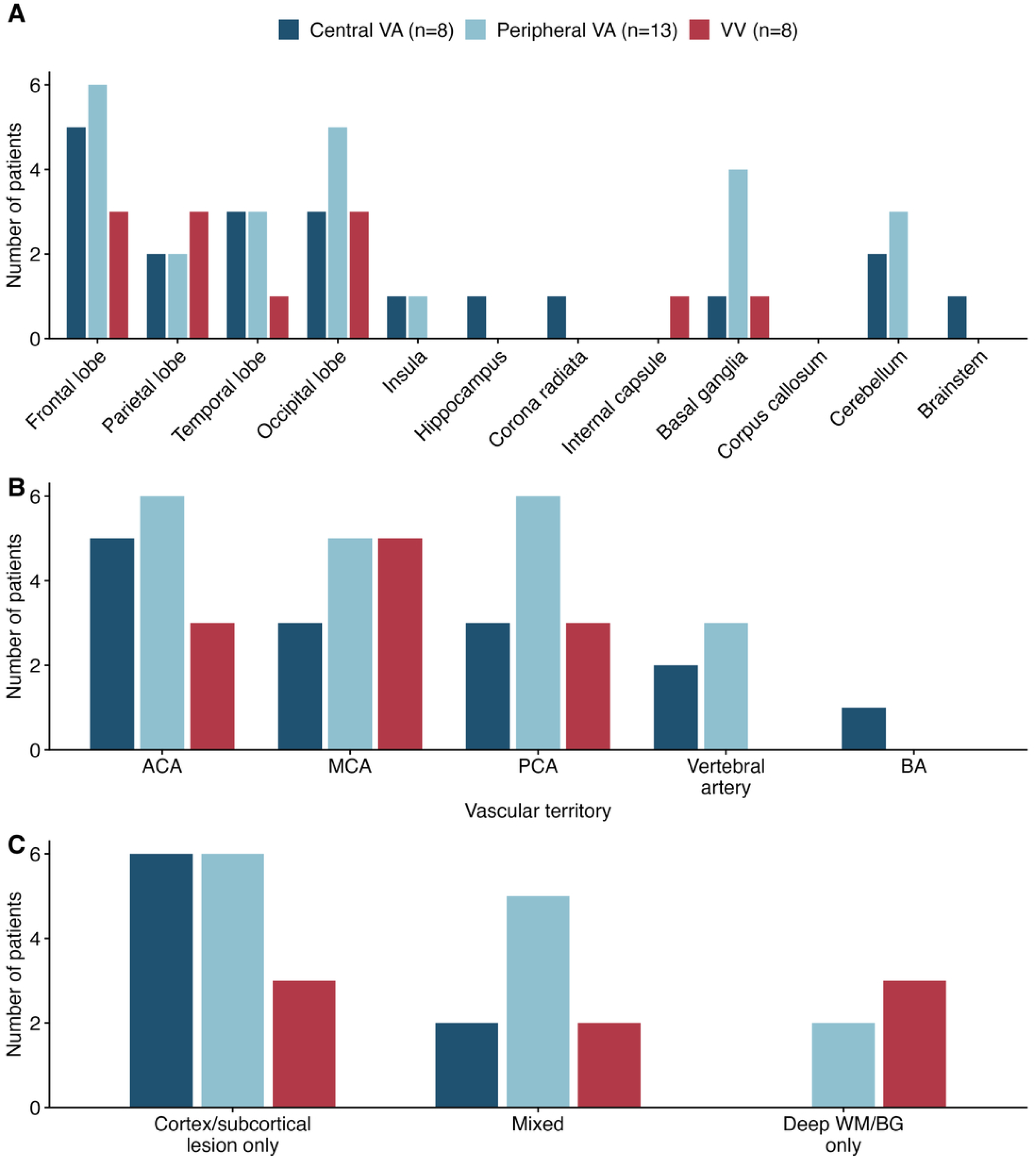
The distribution and topography of acute brain injury. The most common lesion of acute brain injury (ABI) was the frontal lobe, and the parietal, temporal, and occipital lobe and BG were also involved (A). The lesions in anterior circulation, including ACA and MCA, were more often recognized than in posterior circulation (B). Patients receiving central VA-ECMO had at least one cortical/subcortical lesion, while ABI during peripheral VA-ECMO or VV-ECMO involved isolated deep WM/BG lesion (C). Abbreviations: ECMO; Extracorporeal membrane oxygenation VA; Venoarterial, VV; Venoenous, ACA; Anterior cerebral artery, MCA; Middle cerebral artery, PCA; Posterior cerebral artery, BA; Basilar artery, WM; White matter, BG; Basal ganglia.

Central VA-ECMO demonstrated a predominantly cortical/subcortical phenotype, characterized by cortical/subcortical involvement in all patients (100%), and only 25% demonstrating deep WM/BG involvement, while peripheral VA ECMO more commonly led to deep WM/BG involvement (54%) (**Figure 1C**). In VV-ECMO, 62.5% of the patients with ABI had deep WM/BG lesion, suggesting a distinct imaging phenotype compared with central VA-ECMO. Similar findings were observed when analyses were restricted to IS (**Figure S2**).

### Laterality and Bovine Aortic Arch Anatomy

Among VA-ECMO patients with ABI, left hemispheric involvement predominated. Eleven patients (52.4%) had isolated left-sided lesions, seven (33.3%) had bilateral lesions, and only three (14.3%) had isolated right-sided lesions. Compared with central, peripheral VA-ECMO demonstrated a marked left hemispheric predominance (85% left-sided vs. 15% right-sided isolated lesions; *P*=0.002) (**Figure S3**). Among VV-ECMO patients, bilateral lesions were more common (6/8 [75%]), and no patients demonstrated isolated right hemispheric involvement. Aortic arch anatomy was evaluated in 26 patients (VA; 18, VV; 8) with available CT imaging, of whom eight (30.8%) had a bovine arch. Left hemispheric predominance persisted regardless of bovine arch anatomy, with 83.3% of both patients with and without bovine arch demonstrating at least one left-sided lesion in VA-ECMO (**Figure 3**).

**Figure 3:**
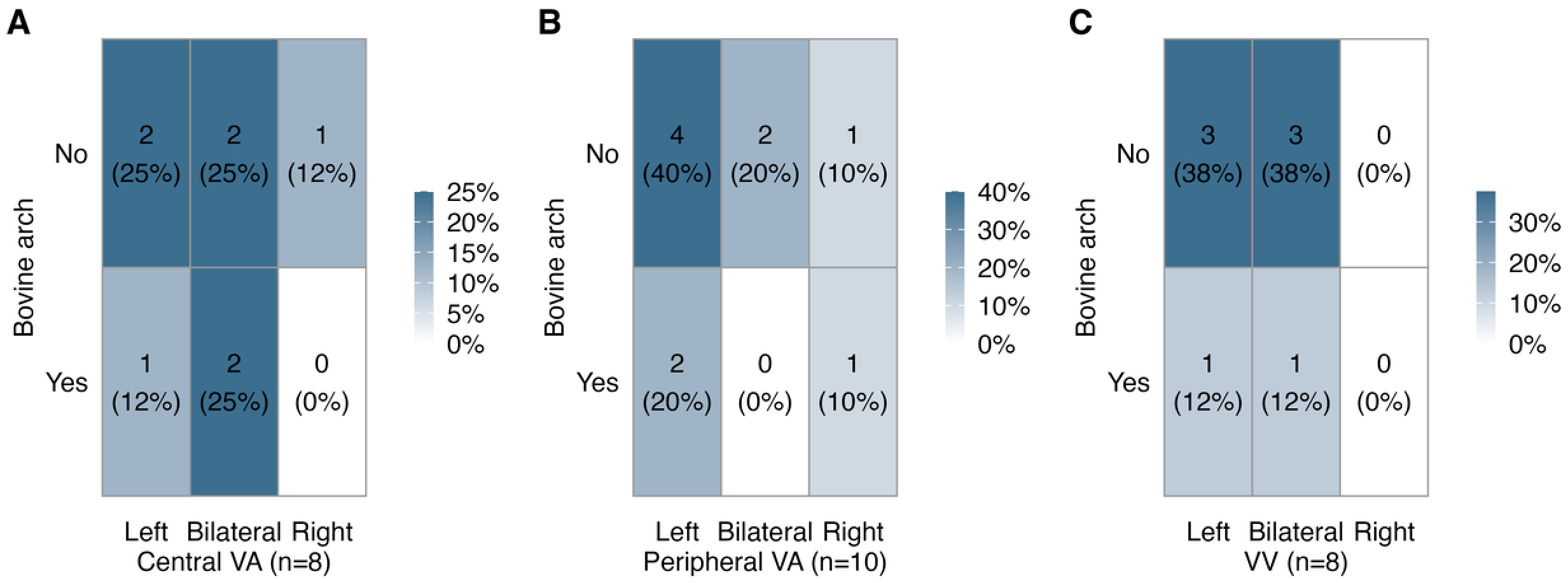
The laterality of acute brain injury and association with bovine arch. The heatmap showed the association between the laterality of acute brain injury (ABI) and the presence of bovine arch in central VA-ECMO (A), peripheral VA-ECMO (B), and VV-ECMO (C). Each cell presents the number of patients and the percentage of the corresponding ECMO mode. Abbreviations: ECMO; Extracorporeal membrane oxygenation, VA; Venoarterial, VV; Venovenous.

### SVD Burden

The prevalence of preexisting cerebral small-vessel disease differed substantially among ECMO configurations. Central VA-ECMO demonstrated the lowest prevalence of SVD (37.5%), whereas SVD was considerably more common in peripheral VA-ECMO (76.9%) and VV-ECMO (75%) (**Figure 4**). Among VV-ECMO, all patients with deep WM or BG lesions had coexisting SVD, compared with 66.7% of patients with only cortical/subcortical lesions, suggesting an association between deep lesion phenotype and preexisting cerebral small-vessel vulnerability.

**Figure 4:**
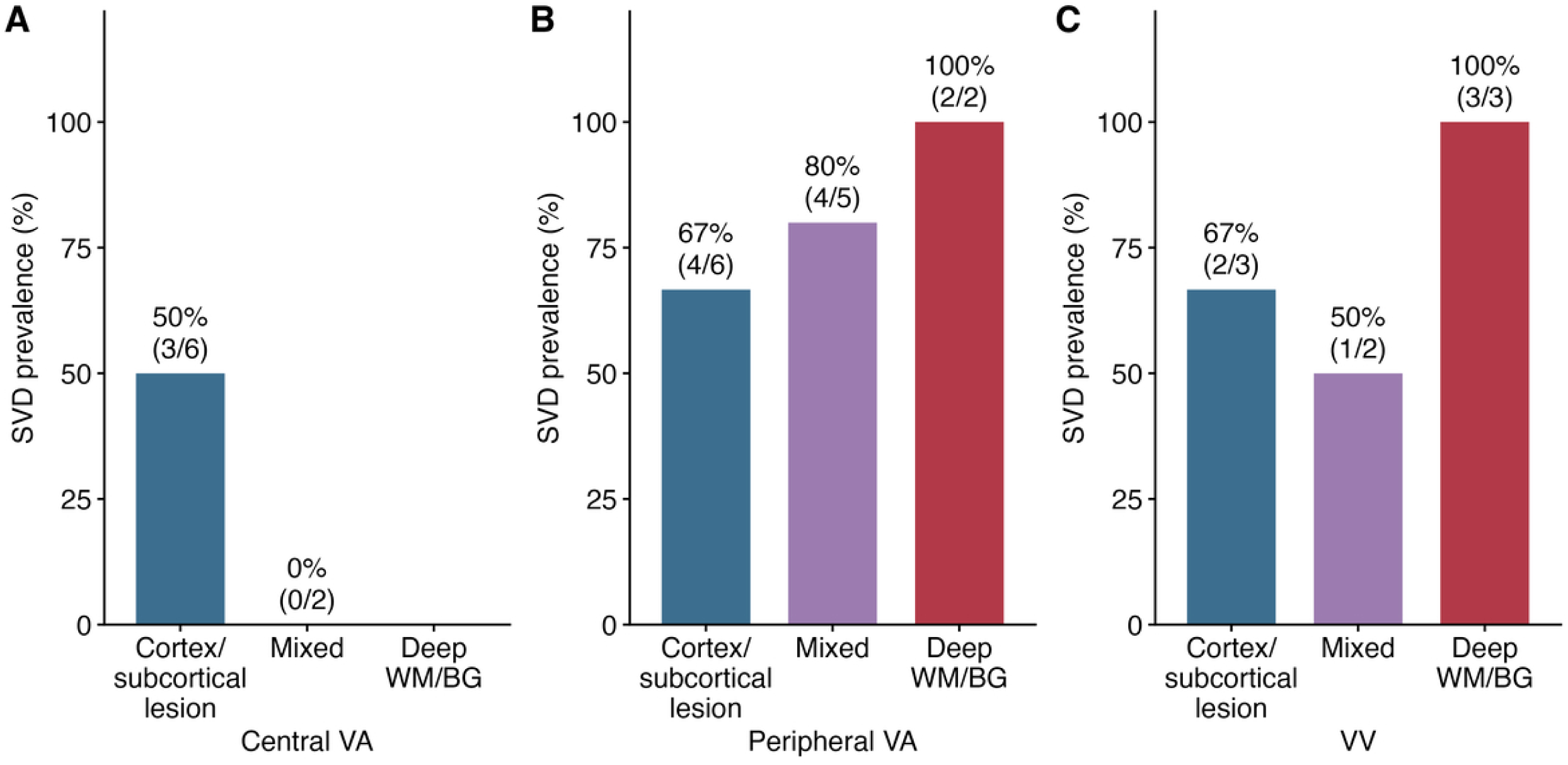
The prevalence of small vessel disease. The prevalence of small vessel disease in central VA-ECMO (A), peripheral VA-ECMO (B) and VV-ECMO (C). Abbreviations: SVD; small vessel disease, ECMO; Extracorporeal membrane oxygenation, VA; Venoarterial, VV; Venovenous. WM; White matter, BG; Basal ganglia.

### Pathophysiological Variables

Among the 25 ABI patients with available echocardiography, PFO (8%), severely reduced ejection fraction (12%), and left ventricular thrombus (0%) were uncommon (**Table S1**). Patients with PFO or reduced ejection fraction demonstrated cortical/subcortical lesions, whereas isolated deep WM or BG lesions occurred in the absence of identifiable cardiac embolic sources. ECMO circuit thrombus was present in 80% of ABI patients.

Among peripheral VA-ECMO patients, those with deep WM or BG lesions demonstrated trends toward higher ECMO pump flow and lower pulse pressure than patients with exclusively cortical/subcortical lesions, although these differences did not reach statistical significance (**Figure 5; Table S2**). Similarly, in VA-ECMO, the higher ECMO pump flow was also recognized in those with deep WM or BG lesions (**Figure S4; Table S3**).

**Figure 5:**
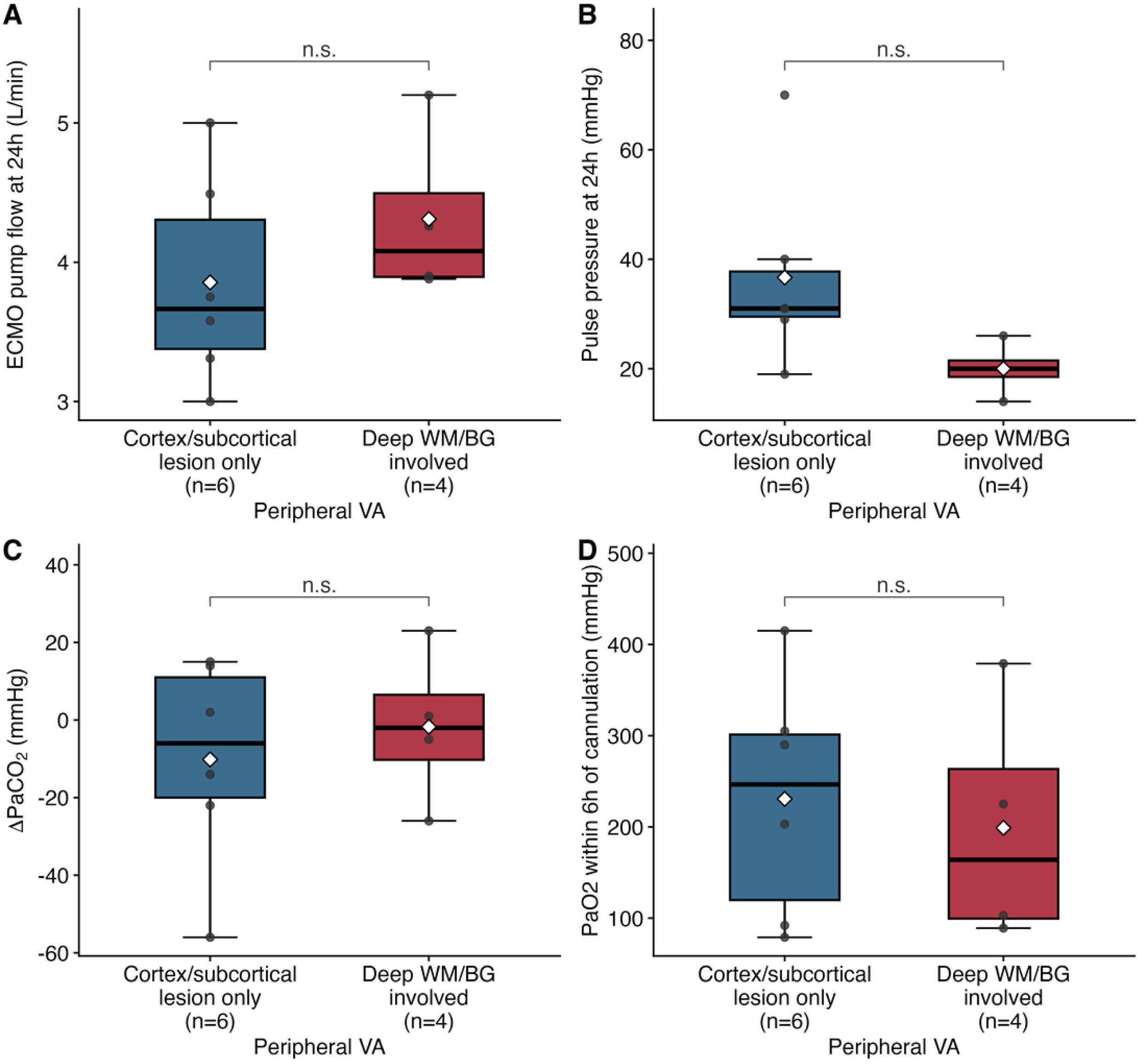
the comparison of physiological parameters between patients with and without deep WM and BG lesion among peripheral VA-ECMO. The comparison of ECMO pump flow (A), pulse pressure 24 hours after cannulation (B), ΔPaCO_2_ (C) and PaO2 within 6 hours (D) between patients with and without deep WM/BG lesion among peripheral VA-ECMO. Abbreviations: PaCO2; Partial arterial carbon dioxide, PaO2; Partial arterial oyxgen, ECMO; Extracorporeal membrane oxygenation, VA; Venoarterial, VV; Venovenous. WM; White matter, BG; Basal ganglia.

## Discussions

We investigated the anatomical distribution of ECMO-associated ABI and characterized its radiological phenotypes in relation to ECMO configuration and pathophysiological findings. Our principal findings were: (1) a marked left hemispheric predominance of ABI during VA-ECMO, particularly during peripheral VA-ECMO; (2) a predominantly cortical/subcortical lesion phenotype in central VA-ECMO; and (3) a higher prevalence of deep WM/BG lesions accompanied by greater preexisting SVD burden in VV-ECMO. These findings suggest that ECMO-associated ABI is heterogeneous and that the underlying mechanisms likely differ according to ECMO configuration. While the arterial cannula in central VA-ECMO is typically inserted into the ascending aorta, providing antegrade blood flow, peripheral VA-ECMO is characterized by retrograde arterial flow from the femoral artery.^23,24^ In contrast, VV-ECMO provides respiratory support without directly altering left ventricular function.^25^ The strength of this study lies not only in the accurate diagnosis and lesion mapping enabled by ULF-pMRI but also in linking distinct imaging phenotypes with ECMO configuration and physiological variables to generate mechanistic hypotheses.

Central VA-ECMO demonstrated a predominantly cortical/subcortical imaging phenotype, consistent with an embolic mechanism. All patients receiving central VA-ECMO had cortical or subcortical lesions, whereas deep WM/BG involvement was uncommon. In addition, ECMO circuit thrombus was frequently identified among ABI patients (80%), whereas other potential cardiac embolic sources, including left ventricular thrombus, severely reduced ejection fraction, and PFO, were uncommon. Our previous study demonstrated cerebral microembolic signals in the middle cerebral artery in 42.2% of patients receiving central VA-ECMO using transcranial Doppler ultrasonography.^10,26^ ECMO circuit thrombosis may both reflect and amplify a systemic prothrombotic state through activation of coagulation and inflammatory pathways and has therefore been proposed as a contributor to embolic ischemic stroke during VA-ECMO.^10,26^ Although our study cannot establish causality, the predominantly cortical lesion distribution together with the high prevalence of ECMO circuit thrombus supports an embolic imaging phenotype in central VA-ECMO.

Peripheral VA-ECMO demonstrated a distinct imaging phenotype characterized by frequent deep WM/BG involvement and marked left hemispheric predominance. More than half of the patients had deep WM/BG lesions, while left-side involvement remained prominent regardless of the presence of bovine aortic arch. Since previous reports have indicated the left hemispheric laterality of embolic stroke in bovine arch and up to 30% of the general population have this variant,^19^ our findings may provide detailed and robust association between the left-side dominance of ABI and peripheral VA-ECMO, regardless of bovine arch. Deep WM/BG lesions are generally associated with cerebral small-vessel pathology, impaired microvascular perfusion, or deep intracerebral hemorrhage, whereas embolic infarction more commonly involves cortical or territorial regions.^27,28^ Furthermore, our previous autopsy study demonstrated cerebral microhemorrhages involving the deep white matter in approximately 60% of patients receiving ECMO.^17^ Together with the observed trend toward higher ECMO pump flow and lower pulse pressure among patients with deep lesions, these findings suggest that altered cerebral hemodynamics, impaired cerebrovascular autoregulation, and cerebral microvascular dysfunction may contribute to this phenotype. At the same time, the persistent left hemispheric predominance suggests that embolic mechanisms and nonphysiological retrograded flow may also contribute, indicating that ABI during peripheral VA-ECMO may represent a mixed phenotype involving both embolic and microvascular mechanisms. These observations should be considered hypothesis-generating and require confirmation in larger studies.

ABI during VV-ECMO demonstrated imaging characteristics distinct from those observed during VA-ECMO. Deep WM/BG lesions were common, and well-established cardiac embolic risk factors, including PFO, were not identified among patients with ABI. Furthermore, patients with ABI demonstrated a greater burden of preexisting SVD than those without ABI. These findings suggest that ABI during VV-ECMO may be more closely related to underlying cerebral small-vessel vulnerability and microvascular dysfunction than to large-artery embolic mechanisms. Because VV-ECMO preserves native cardiac output and does not directly alter systemic arterial flow, these observations further support the concept that different ECMO configurations may produce distinct imaging phenotypes through different pathophysiological pathways.

Our study has several limitations. First, the sample size was relatively small, and all mechanistic associations should be considered hypothesis-generating. Nevertheless, this represents the first study to systematically characterize acute brain injury using ULF-pMRI in patients receiving ECMO, a population in whom conventional MRI is generally not feasible. Given the technical challenges of bedside neuroimaging during ECMO, these findings provide important preliminary insights into the radiological phenotypes and potential mechanisms of ECMO-associated ABI and establish a foundation for future multicenter validation studies. Second, the timing of ULF-pMRI was not standardized because imaging was performed according to each patient’s clinical condition and logistical considerations in the intensive care unit. Consequently, lesion evolution may have differed among patients at the time of imaging. Third, ULF-pMRI may lead to underdiagnosis of brainstem and cerebellum lesions because of susceptibility to artifacts,^29^ and the ABI involving those regions may be underestimated. Lastly, the observational design precludes causal inference between the identified imaging phenotypes and the proposed pathophysiological mechanisms.

## Conclusions

ULF-pMRI identified distinct imaging phenotypes of ECMO-associated ABI that differed by ECMO configuration. Central VA-ECMO demonstrated a predominantly cortical/subcortical lesion pattern consistent with an embolic phenotype, whereas peripheral VA-ECMO and VV-ECMO more frequently exhibited deep WM and BG involvement, suggesting contributions from cerebral small-vessel vulnerability or microvascular dysfunction. As the first ULF-pMRI study of ABI during ECMO, these findings provide a framework for imaging-based phenotyping and generate mechanistic hypotheses that warrant validation in larger prospective multicenter studies.

## Data Availability

per request, we will review it.

## Acknowledgement

The Hopkins Exploration, Research, and Advancement in Life support Devices (HERALD) group collaborators are as follows: Daniel Brodie, Marc Sussman, Diane Alejo, Trish Brown, Karlo Capili, Zachary Darby, Harry Flaster, Andrew Geeza, Jin Kook Kang, Shivalika Khanduja, Madison Malfitano, Bo Soo Kim, Daniel Forsman, Scott Stephens, Steve Keller, Errol Bush, Pedro Alejandro Mendez-Tellez, Jaeho Hwang, Nitish Thakor, Xiaoyan Zhu, Christopher Wilcox, Andrew Kalra, Leon Fan, Amy Feng, Chang Liu, Winnie Liu, Ernesto Marin, Alessandro Ascani Orsini, Pierce Logan Perkins, Beichen Shen, Sukethram Sivakumar, Tony Zhu, Jayden Allegakoen, Yipeng Wang, Joshua Russman, Kayoung Kwon, Jeena Lee.

## Author contribution

Kotaro Noda contributed to methodology development, data acquisition and drafting of the manuscript. Yaman B. Ahmed contributed to study conceptualization, methodology development, and data acquisition. Hyun Yi Woo contributed to editing of the manuscript, critically reviewed the manuscript. Shivalika Khanduja contributed to study conceptualization, methodology development, and data acquisition. Yipeng W contributed to data acquisition. Zoe Soulé contributed to data acquisition. Siyu Wang contributed to editing of the manuscript, critically reviewed the manuscript. Madison Malfitano contributed to editing the manuscript and critical review of the manuscript. Bo Soo Kim contributed to study conceptualization, methodology development, and data acquisition. Christopher Wilcox contributed to study conceptualization, methodology development, and data acquisition. Glenn Whitman contributed to resource provision, study support, funding acquisition, and project supervision. Sung-Min Cho oversaw study conceptualization, funding acquisition, project supervision, critical revision of all manuscript drafts, and final approval of the manuscript.

## Abbreviations

ABI: acute brain injury
ACA: anterior cerebral artery
ANOVA: analysis of variance
BA: basilar artery
BG: basal ganglia
CHF: chronic heart failure
CKD: chronic kidney disease
CT: computed tomography
DSWMH: deep and subcortical white matter hyperintensity
ECMO: extracorporeal membrane oxygenation
EF: ejection fraction
HERALD: Hopkins Exploration, Research, and Advancement in Life support Devices
HIBI: hypoxic-ischemic brain injury
ICH: intracranial hemorrhage
IPH: intraparenchymal hemorrhage
IQR: interquartile range
IS: ischemic stroke
MCA: middle cerebral artery
MRI: magnetic resonance imaging
PaCO2: partial pressure of arterial carbon dioxide
PCA: posterior cerebral artery
PFO: patent foramen ovale
PVH: periventricular hyperintensity
SAH: subarachnoid hemorrhage
SD: standard deviation
SDH: subdural hematoma
SVD: small-vessel disease
ULF-pMRI: ultra-low-field portable magnetic resonance imaging
VA: venoarterial (ECMO configuration) or vertebral artery (vascular territory)
VV: venovenous
WM: white matter
ΔPaCO2: change in arterial carbon dioxide

